# Social Inequalities in Suicide Across Argentine Provinces: Trends Before and After COVID-19 Pandemic

**DOI:** 10.1101/2025.04.25.25326449

**Authors:** Carlos M. Leveau

**Affiliations:** Centro de Salud Mental Comunitaria “Mauricio Goldenberg”, Universidad Nacional de Lanús, Argentina; Consejo Nacional de Investigaciones Científicas y Técnicas (CONICET), Argentina

**Keywords:** Health Inequities, Socioeconomic Factors, Pandemics, Educational Status, Latin America

## Abstract

**Background:** Few studies have examined social inequalities in suicide by comparing the COVID-19 pandemic with previous years. These studies, largely concentrated on high-income countries, have produced mixed results and have not included post-pandemic years in their analyses. Our objective was to analyze social inequalities in suicide across Argentine provinces during the period 2015–2023.

**Methods:** This is a retrospective quantitative study based on secondary data analysis. We employed Poisson regressions to model the temporal trends in suicide rates by educational level in the provinces of Jujuy, Mendoza, and San Juan from 2015 to 2023. Additionally, we calculated suicide rate ratios to examine temporal variations in relative inequalities by educational level.

**Results:** Compared to those with medium-high education, individuals with low education— across sexes, men, and the 25–44 age group—had higher suicide risk in all years except 2021. During the pandemic, women’s educational mortality ratios fell below 1, contrasting with pre-pandemic (2015–2019) and post-pandemic (2022–2023) periods. For men, suicide rate ratios exceeded 1 in all periods but were smaller during and after the pandemic. The 25–44 age group mirrored men’s pattern, while the 45+ group showed no educational inequalities in suicide during and after the pandemic.

**Conclusions:** The COVID-19 pandemic appeared to temporarily narrow the social suicide gap, though variations by sex and age were observed. This potential effect of the pandemic seemed to be short-lived, with the 2022–2023 period showing a return to pre-pandemic levels of social inequalities in suicide.

**Key messages:** *What is already known on this topic:* Social inequalities in suicide exist, although the evidence in Latin America remains limited and inconclusive.

*What this study adds:* We examined social inequalities in suicide across three periods: pre-pandemic, during the COVID-19 pandemic, and post-pandemic. The suicide gap between the least educated and the most educated populations tended to narrow during the pandemic years (2020–2021), with a particularly pronounced decline observed among women with low educational attainment.

*How might this study affect research, practice, or policy:* Further in-depth studies, incorporating qualitative approaches, are necessary to confirm the potential positive impact of social policies on the well-being of women with low socioeconomic status during the COVID-19 pandemic.

## Introduction

The COVID-19 pandemic caused an abrupt drop in economic activity, massive social isolation, and uncertainty and fear of contagion from the new SARS-CoV-2 virus. In Latin American societies, where a significant proportion of the adult population subsists in the informal economy and chronic job insecurity, the COVID-19 pandemic – and the quarantine measures implemented – left these populations in a sudden situation of total helplessness. Therefore, a widening of the social gap in suicide would be expected with the onset of the pandemic in Latin America.

Although an increase in suicides was predicted at the beginning of the COVID-19 pandemic^1^, associated with quarantine measures, their impact on the economy, and the uncertainty generated by the increase in infections and deaths, empirical studies have mostly found stability, or even a decrease, in suicides during the first year of the pandemic^2–10^. However, this general stability of suicides could mask social inequalities, since stay-at-home measures would protect populations with a high socioeconomic level to a greater extent –for example, through remote work– and populations with a low socioeconomic level to a lesser extent –face-to-face jobs in precarious conditions.

Despite the studies that have studied suicide trends and their association with the COVID-19 pandemic, few studies have analysed its socioeconomic inequalities and its post-pandemic trend. Changes in social inequalities in suicide following the COVID-19 pandemic have been studied in only a few high-income countries. Three studies conducted in the United States showed an increase in suicides in non-White populations during the quarantine months, while suicides in non-Hispanic white populations decreased^11–13^. In contrast, a study conducted in the state of Michigan found no differences between white and black populations before and after the pandemic^14^. For three states in Australia, suicides associated with financial problems and unemployment did not increase during the months of the COVID-19 pandemic^15^. Similarly, in Sweden the pandemic had no impact on differences in suicide by educational level^16^.

Therefore, evidence is lacking in low- and middle-income countries, where cultural factors might modify the social gradient of suicide found in high-income countries^17^. Although low socioeconomic level populations have a higher risk of suicide in Argentina, the social gap in suicide tended to decrease during the severe economic crisis of 1999-2002^18^. Thus, our objective was to analyse the social inequalities of suicide in Argentine provinces, during the period 2015- 2023.

## Methods

This is a retrospective quantitative study based on secondary data analysis. Data on suicide deaths (ICD-10 codes: X60–X84) from 2015 to 2023 were obtained from the national vital statistics provided by the Ministry of Health. These records were disaggregated by age, sex, province of residence, and educational level. Additionally, suicides were categorized based on residence in the capital city of each province (main metropolitan area) versus the rest of the province. Due to small numbers of suicide cases, educational level was dichotomized into two groups: low (less than complete secondary education) and medium-high (completed secondary education or higher). Age was treated as a categorical variable in the regression models, with the following groups: 25–34, 35–44, 45–54, 55–64, and 65 years or older.

The study population included three Argentine provinces: Jujuy, Mendoza, and San Juan. These provinces were selected because, during the period 2015–2023, 90% or more of annual suicide fatalities included information on the educational level of the deceased (Table 1A, Supplementary Material). Together, these provinces are home to 3,659,582 inhabitants, representing approximately 8% of Argentina’s total population. Population estimates for the period 2015–2023, disaggregated by age, sex, province of residence, metropolitan status, and educational level, were derived through linear projections using data from the 2010 and 2022 population censuses.

Due to overdispersion in the data, negative binomial regression models were employed to examine the relationship between educational level and suicide rates, stratified by sex and three age groups (25–44 years, 45–64 years, and 65 years and older). The explanatory variables included sex, age, educational level, province of residence, metropolitan status, and year. The deceased’s educational level was used as a proxy for socioeconomic status, as it is the only variable of this kind available in vital statistics. Furthermore, numerous studies have demonstrated a strong correlation between educational attainment and various measures of socioeconomic status, such as income, wealth, and overall quality of life^19,20^.

The dependent variables were counts of suicides for the following groups: (i) all individuals aged 25 and over; (ii) males aged 25 and over; (iii) females aged 25 and over; (iv) individuals aged 25–44; (v) individuals aged 45–64; and (vi) individuals aged 65 and over. Mortality ratios were calculated by comparing suicide rates in the low-education population to those in the medium-high education population, which served as the reference group.

For each dependent variable, we estimated three models. The first model assessed the relationship between educational level and suicide mortality, adjusting for sex, age, province of residence, metropolitan status, and year. The second model introduced an interaction term between year and educational level to evaluate the differential trends in mortality rates and mortality ratios by educational level over time. The third model incorporated a categorical variable representing three subperiods: pre-pandemic (2015–2019), pandemic (2020–2021), and post-pandemic (2022–2023). An interaction term between this subperiod variable and educational level was included to compare mortality rates and mortality ratios across educational levels and subperiods. Although there was a final peak in COVID-19 deaths between January and February 2022, mass vaccination against the SARS-CoV-2 virus had already been achieved this year^21^. To avoid collinearity, the year variable was excluded from this model. Marginal predicted rates were calculated to estimate mortality rates and mortality ratios by educational level for each year (second model) and across subperiods (third model).

To assess the robustness of our findings in light of potential under-reporting of suicides, all models were recalculated by including deaths of undetermined intent by hanging (ICD-10 code: Y20) and drowning (ICD-10 code: Y21) in the suicide counts. These mechanisms of death due to undetermined intent (hanging, strangulation, suffocation, drowning, and submersion) were included in the analysis because there is evidence suggesting a higher likelihood of these cases being misclassified suicides^22^. However, undetermined injuries without a specified mechanism of death (ICD-10 code: Y34) were excluded from the analysis, as the lack of detailed information makes it difficult to attribute them to a specific type of injury or to reliably classify them as suicides^23^. All analyses were conducted using Stata version 13.1 (StataCorp, College Station, TX).

## Results

Between 2015 and 2022, a total of 1,924 suicides were recorded among individuals aged 25 and older residing in the provinces of Jujuy, Mendoza, and San Juan. Of these, 1,890 cases (98%) included complete data on age, sex, educational level, and department of residence. Table 2A (Supplementary Material) presents the distribution of suicides by year of occurrence, educational level, sex, age, and place of residence. In Mendoza and San Juan, suicide frequencies declined in 2020 but increased during 2021–2023 (Table A2, Supplementary Material). Across all three provinces, suicide frequencies were higher among populations with lower educational attainment, with the highest proportion observed in Jujuy. Notably, Jujuy also exhibited the youngest age profile of suicide in absolute numbers (Table A2, Supplementary Material). Regarding sex, a higher percentage of suicides occurred among men, with similar proportions across the three provinces. Finally, while Jujuy reported a higher percentage of suicides outside the metropolitan area of the provincial capital, the opposite pattern was observed in Mendoza and San Juan (Table A2, Supplementary Material).

Considering the entire period 2015-2023 as a whole, in both men and women, and in the three age groups, the risk of suicide was higher in the population with a low educational level (up to incomplete secondary education) compared to the population with a higher educational level (Table 1). Men and the young adult population (25-44 years) showed the highest mortality ratios.

**Table 1.**
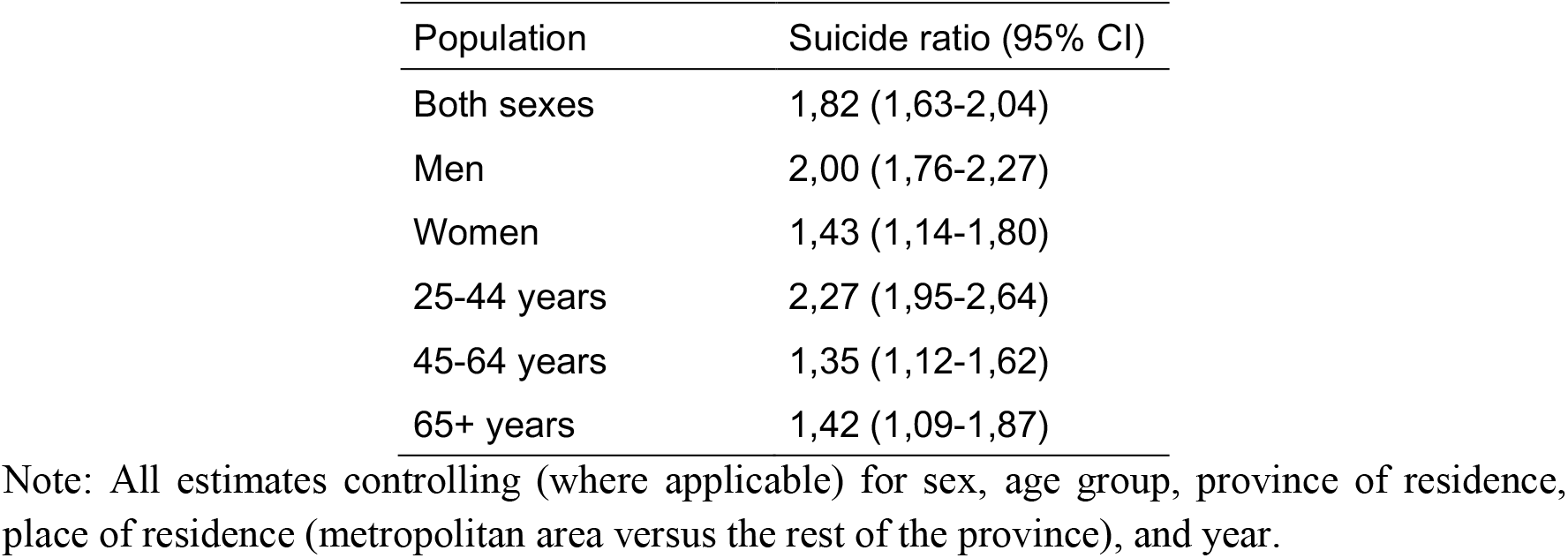
Suicide ratios (95% confidence intervals, CIs) for the low educational level population (using medium-high educational level population as the standard) in three Argentinian provinces (Jujuy, Mendoza, and San Juan), 2015-2023.

| Population | Suicide ratio (95% CI) |
| --- | --- |
| Both sexes | 1,82 (1,63-2,04) |
| Men | 2,00 (1,76-2,27) |
| Women | 1,43 (1,14-1,80) |
| 25-44 years | 2,27 (1,95-2,64) |
| 45-64 years | 1,35 (1,12-1,62) |
| 65+ years | 1,42 (1,09-1,87) |

Note: All estimates controlling (where applicable) for sex, age group, province of residence, place of residence (metropolitan area versus the rest of the province), and year.

Figure 1 shows the temporal trends in suicide rates by educational level. While suicide rates among individuals aged 25 and older with low educational attainment declined to their lowest point in 2021, rates among those with medium-high educational attainment peaked in the same year (Figure 1.A).This convergence of suicide rates by educational level in 2021 seems to be largely explained by the combination of maximum peaks in men and the population aged 25-64 years (figures 1.B, 1.D and 1.E), and the abrupt decline in rates in women with low educational levels (Figure 1.C).

**Figure 1.**
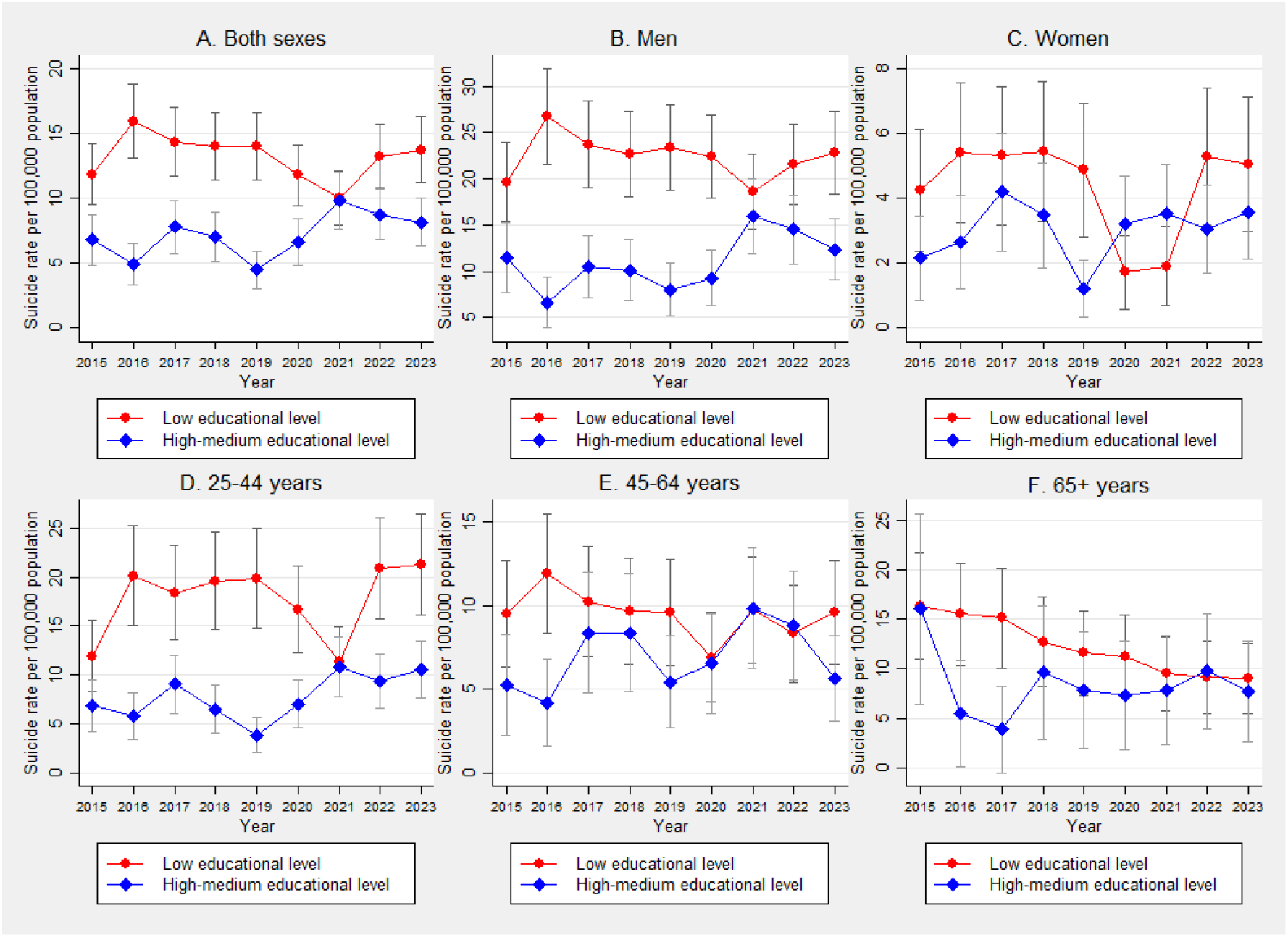
Annual suicide rate per 100,000 population aged 25 years and over in the population of three Argentinian provinces classified by educational level. Vertical segments are 95% confidence intervals. Note: All estimates controlling (where applicable) for sex, age group, province of residence, and place of residence (metropolitan area versus the rest of the province).

Compared to the population with a medium-high educational level, the population with a low educational level of both sexes, men, and the population aged 25-44 showed a higher risk of suicide in all years, except in 2021 (Figure 2). Although confidence intervals for educational mortality ratios in women included 1 in the annual analysis (Figure 2), these intervals were less than 1 considering the three periods –pre-pandemic, pandemic, and post-pandemic (Figure 3). For men, suicide rate ratios exceeded 1 in all three periods, though these relative inequalities were smaller during the pandemic and post-pandemic periods (Figure 3). Regarding age groups, the 25–44 age group exhibited a similar pattern to men, while the 45+ age group showed no educational inequalities in suicide during the pandemic and post-pandemic periods (Figure 3). Robustness checks that included deaths from injuries of undetermined intent involving hanging or drowning (codes Y20–Y21) produced results highly consistent with analyses restricted to confirmed suicides (Table 3A and Figures 1A–3A, Supplementary Material).

**Figure 2.**
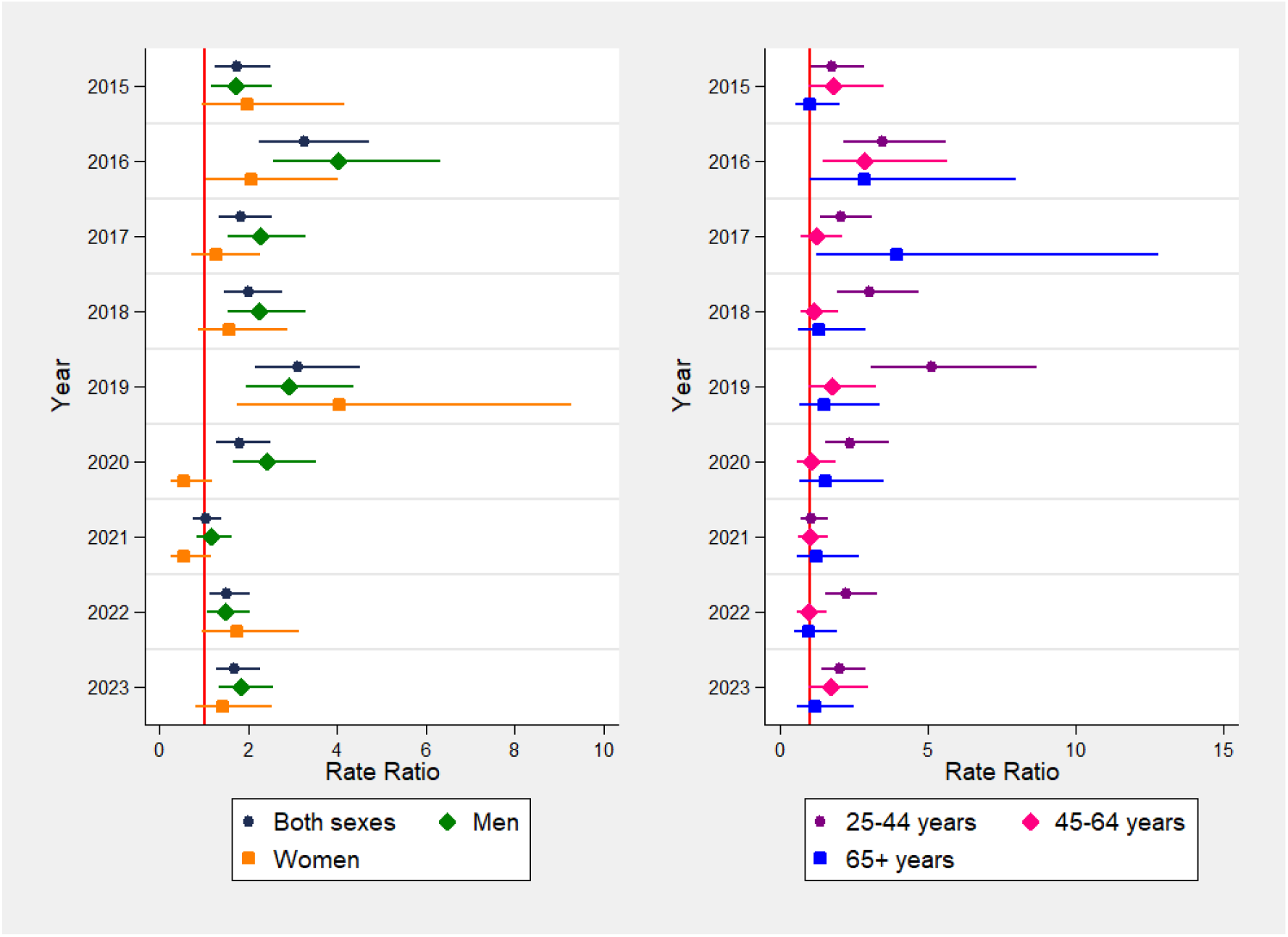
Annual suicide rate ratios using as standard the population of medium-high educational level. Vertical segments are 95% confidence intervals for the ratio of mortality rates computed with mortality including all explanatory variables and an interaction between year and educational level. Note: All estimates controlling (where applicable) for sex, age group, province of residence, and place of residence (metropolitan area versus the rest of the province).

**Figure 3.**
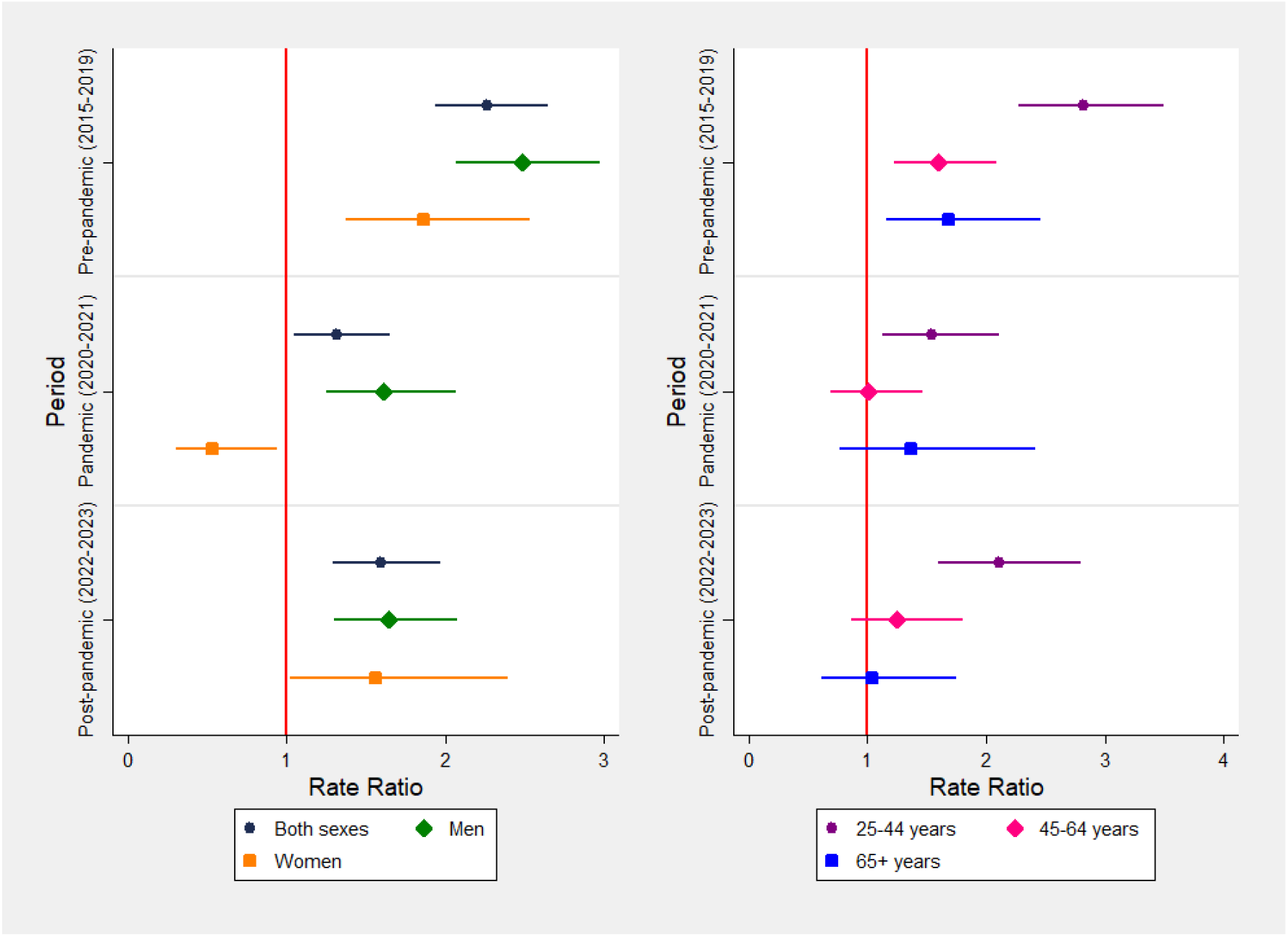
Suicide rate ratios using as standard the population of medium-high educational level. Vertical segments are 95% confidence intervals for the ratio of mortality rates computed with mortality including all explanatory variables and an interaction between period and educational level. Note: All estimates controlling (where applicable) for sex, age group, province of residence, and place of residence (metropolitan area versus the rest of the province).

## Discussion

We studied educational inequalities in suicide in three provinces of Argentina during 2015- 2023. We found a decrease in relative educational inequalities in suicide during 2020-2021 compared to 2015-2019, although populations with low educational attainment had a higher risk of suicide throughout 2015-2023. These trends were replicated in men and the population aged 25-44. In contrast, women with low educational attainment showed a lower risk of suicide during the 2020-2021 period, although this population again had a higher risk of suicide during 2022-2023.

Our finding of a decrease in social inequalities in suicide during 2020-2021 is not consistent with studies conducted in the United States^11–13^. Our findings in the Argentine provinces may be linked to the social policies implemented by the National Government immediately following the introduction of population mobility restrictions. Starting in April 2020, the National Government implemented the Emergency Family Income (IFE), planned to reach households with unemployment, informal employment, low-income self-employed workers, and beneficiaries of social assistance^24^. There were three IFE payments between April and September 2020, reaching almost 9 million people^24^. This economic aid to households with a low socioeconomic level could have reduced the level of uncertainty and vulnerability in these populations, which could explain the decrease in suicide rates during 2020-2021. In addition, for those companies with difficulties in paying salaries, the National Government implemented the Emergency Assistance Program for Work and Production (ATP), with the aim of sustaining employment in the economic activities most severely affected during the first months of the COVID-19 pandemic ^25^. Although IFE and ATP ended at the end of 2020, more targeted economic aid for companies with low demand problems continued from 2021^26^.

Suicide rates in women with low educational levels showed a marked decline in both 2020 and 2021. Related to this, the IFE had a greater impact on women under high social vulnerability: 55% of IFE beneficiaries were women, reaching 59% of women in the population aged 35-44^24^. In addition, young mothers of low socioeconomic status, who were previously banked by the National Government when receiving other social assistance, were quickly reached by the IFE during the start of the quarantine measures^24^.

In this context, it can be added that women of low socioeconomic status (SES) tend to have more children than their medium- and high-SES counterparts, a factor that may offer greater protection against suicide^27^, especially at a time of high social isolation such as the COVID-19 pandemic. In Brazil, a massive income transfer program to low socioeconomic populations was associated with a lower incidence of suicide, with a stronger association in women and the population aged 25-59 years^28^.

At the same time, there was an increase in suicide rates in the population of medium-high educational level. This increase is not expected according to some studies carried out in the United States^12,13^. This increase, observed in Argentine provinces, peaked in 2021 among men and individuals aged 45–64. Those self-employed workers with medium incomes were not covered by the state aid of the IFE during 2020, potentially leaving them in a highly vulnerable position, particularly those aged 45–64. Then, in 2021, despite the initiation of COVID-19 vaccinations, the deadliest wave of the pandemic occurred, disproportionately affecting individuals under 60 compared to the first wave. This likely heightened uncertainty for those re-entering the labour market amid the reopening of economic activities, particularly men aged 45– 64, who faced a higher risk of COVID-19 mortality during the 2021 wave compared to the initial wave in 2020.

Furthermore, the closure of commercial economic activities deemed non-essential during the quarantine may have disproportionately affected adult men with medium-to-high educational attainment. As Girard^29^ highlights, job loss in adulthood can severely threaten an individual’s self-concept, particularly when re-entering the workforce becomes more difficult with age. This risk may be especially acute among high-skilled professionals, whose careers are often defined by achievement-based progression and a strong reliance on occupational identity.

It was hypothesized that the socially unequal impact of COVID-19 infection and mortality would correspond with the social inequalities in suicide rates^13^. In two of the provinces in our study area, COVID-19 mortality disproportionately impacted the population with a low educational level. However, this was not associated with an increase in educational inequalities in suicide, especially in the older adult population (65+ years), given that the loss of a partner could be a factor associated with suicide^30^.

There is concern about the worsening mental health of the population following the COVID-19 pandemic^31^. Furthermore, the social gap in mental illness is expected to increase in the post-pandemic era^31^. Although our results show an increase in relative social inequalities in suicide during 2022-2023, these were not higher than those recorded in the pre-pandemic period 2015- 2019.

This study has several limitations that should be acknowledged. First, the analysis focused on three Argentine provinces limits the generalizability of the findings to the rest of the country. This is particularly relevant given the potential influence of province-specific social policies implemented in response to the COVID-19 pandemic, which may have affected suicide rates. Second, the reliance on linear population projections may underestimate interannual variations in at-risk populations. For instance, the closure of universities in metropolitan areas during 2020 could have prompted short-term migration of young people to other parts of the province. However, data on the existence and magnitude of such population movements are unavailable, limiting our ability to account for these dynamics in the analysis. Third, the 2022 Census did not include questions on the marital status of the adult population, a variable frequently linked to suicide risk^32–35^, limiting the ability to explore this potential association in the analysis. Fourth, unfortunately, the month of death was not available in vital statistics databases. This information would have made it possible to analyse the variations in social inequalities in suicide considering the pandemic period from March 2020, when the first cases of COVID-19 were recorded in Argentina.

The COVID-19 pandemic appeared to temporarily narrow the social suicide gap, though variations by sex and age were observed. This potential effect of the pandemic seemed to be short-lived, with the 2022–2023 period showing a return to pre-pandemic levels of social inequalities in suicide. The absence of social inequalities in suicide among women during the COVID-19 pandemic, linked to a decrease in suicide rates among women with low educational attainment, underscores the need for transdisciplinary studies to assess the role of national cash transfer policies targeting the most vulnerable populations during the pandemic.

## Data Availability

All data produced are available online at https://www.datos.gob.ar/

## Statements and Declarations

### Consent for publication

Not applicable.

### Ethics approval and consent to participate

Not applicable.

## 1. Supplementary Material

**Table 1A.** Percentage of total suicides with educational level data, 2015–2023.

| Year | Jujuy |  | Mendoza |  | San Juan |  |
| --- | --- | --- | --- | --- | --- | --- |
|  | Total count of suicides* | Percentage of total suicides with educational level | Total count of suicides* | Percentage of total suicides with educational level | Total count of suicides* | Percentage of total suicides with educational level |
| 2015 | 37 | 100,0 | 117 | 98,3 | 23 | 100,0 |
| 2016 | 59 | 100,0 | 118 | 100,0 | 27 | 100,0 |
| 2017 | 33 | 100,0 | 147 | 99,3 | 41 | 100,0 |
| 2018 | 43 | 100,0 | 136 | 100,0 | 35 | 94,3 |
| 2019 | 53 | 98,1 | 100 | 97,0 | 40 | 100,0 |
| 2020 | 72 | 100,0 | 94 | 95,7 | 31 | 96,8 |
| 2021 | 45 | 100,0 | 120 | 96,7 | 52 | 94,2 |
| 2022 | 52 | 100,0 | 138 | 97,8 | 57 | 98,2 |
| 2023 | 66 | 98,5 | 129 | 98,4 | 59 | 96,6 |
| Totals | 460 |  | 1099 |  | 365 |  |

\* Suicides of 25+ years.

**Table 2A.** Characteristics of suicides in the Argentine provinces of Jujuy, Mendoza, and San Juan (2015-2023). Percentages in parentheses.

|  | Jujuy | Mendoza | San Juan |
| --- | --- | --- | --- |
| *Year (%)* |  |  |  |
| 2015 | 37 (8,2) | 115 (10,6) | 23 (6,5) |
| 2016 | 58 (12,8) | 118 (10,9) | 27 (7,6) |
| 2017 | 33 (7,3) | 146 (13,5) | 41 (11,5) |
| 2018 | 42 (9,3) | 136 (12,6) | 33 (9,3) |
| 2019 | 52 (11,5) | 100 (9,2) | 40 (11,2) |
| 2020 | 71 (15,7) | 89 (8,2) | 30 (8,4) |
| 2021 | 44 (9,7) | 116 (10,7) | 49 (13,8) |
| 2022 | 52 (11,5) | 135 (12,5) | 56 (15,7) |
| 2023 | 63 (13,9) | 127 (11,7) | 57 (16,0) |
| Totals | 452 (100) | 1082 (100) | 356 (100) |
| *Educational Level (%)* |  |  |  |
| Low | 329 (72,8) | 709 (65,5) | 237 (66,6) |
| Medium-high | 123 (27,2) | 373 (34,5) | 119 (33,4) |
| Totals | 452 (100) | 1082 (100) | 356 (100) |
| *Sex (%)* |  |  |  |
| Female | 83 (18,4) | 244 (22,6) | 57 (16,0) |
| Male | 369 (81,6) | 838 (77,4) | 299 (84,0) |
| Totals | 452 (100) | 1082 (100) | 356 (100) |
| *Age (%)* |  |  |  |
| 25-34 | 187 (41,4) | 312 (28,8) | 125 (35,1) |
| 35-44 | 109 (24,1) | 238 (22,0) | 77 (21,6) |
| 45-54 | 51 (11,3) | 170 (15,7) | 62 (17,4) |
| 55-64 | 44 (9,7) | 131 (12,1) | 40 (11,2) |
| 65+ | 61 (13,5) | 231 (21,3) | 52 (14,6) |
| Totals | 452 (100) | 1082 (100) | 356 (100) |
| *Metropolitan area (%)* |  |  |  |
| Yes | 210 (46,5) | 655 (60,5) | 280 (78,7) |
| No | 242 (53,5) | 427 (39,5) | 76 (21,3) |
| Totals | 452 (100) | 1082 (100) | 356 (100) |

**Table 3A.** Suicide* ratios (95% confidence intervals, CIs) for the low educational level population (using medium-high educational level population as the standard) in three Argentinian provinces (Jujuy, Mendoza, and San Juan), 2015-2023.

| Population | Suicide ratio (95% CI) |
| --- | --- |
| Both sexes | 1,85 (1,66-2,07) |
| Men | 2,04 (1,80-2,31) |
| Women | 1,45 (1,16-1,81) |
| 25-44 years | 2,29 (1,97-2,66) |
| 45-64 years | 1,40 (1,17-1,68) |
| 65+ years | 1,43 (1,10-1,87) |

\* Suicides including deaths of undetermined intent by hanging (ICD-10 code: Y20) and drowning (ICD-10 code: Y21).

**Figure 1A.**
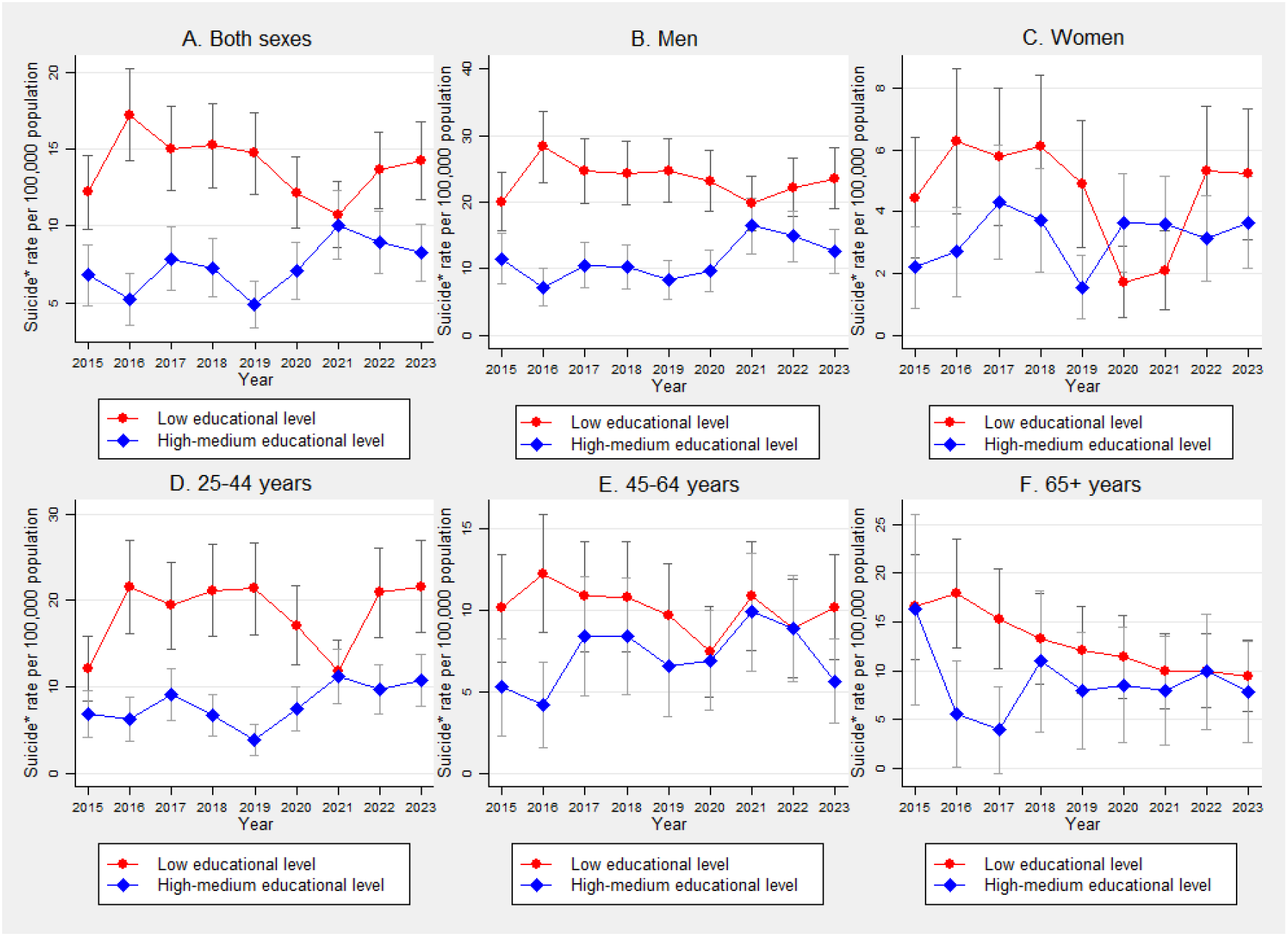
Annual suicide* rate per 100,000 population aged 25 years and over in the population of three Argentinian provinces classified by educational level. Vertical segments are 95% confidence intervals. * Suicides including deaths of undetermined intent by hanging (ICD-10 code: Y20) and drowning (ICD-10 code: Y21).

**Figure 2A.**
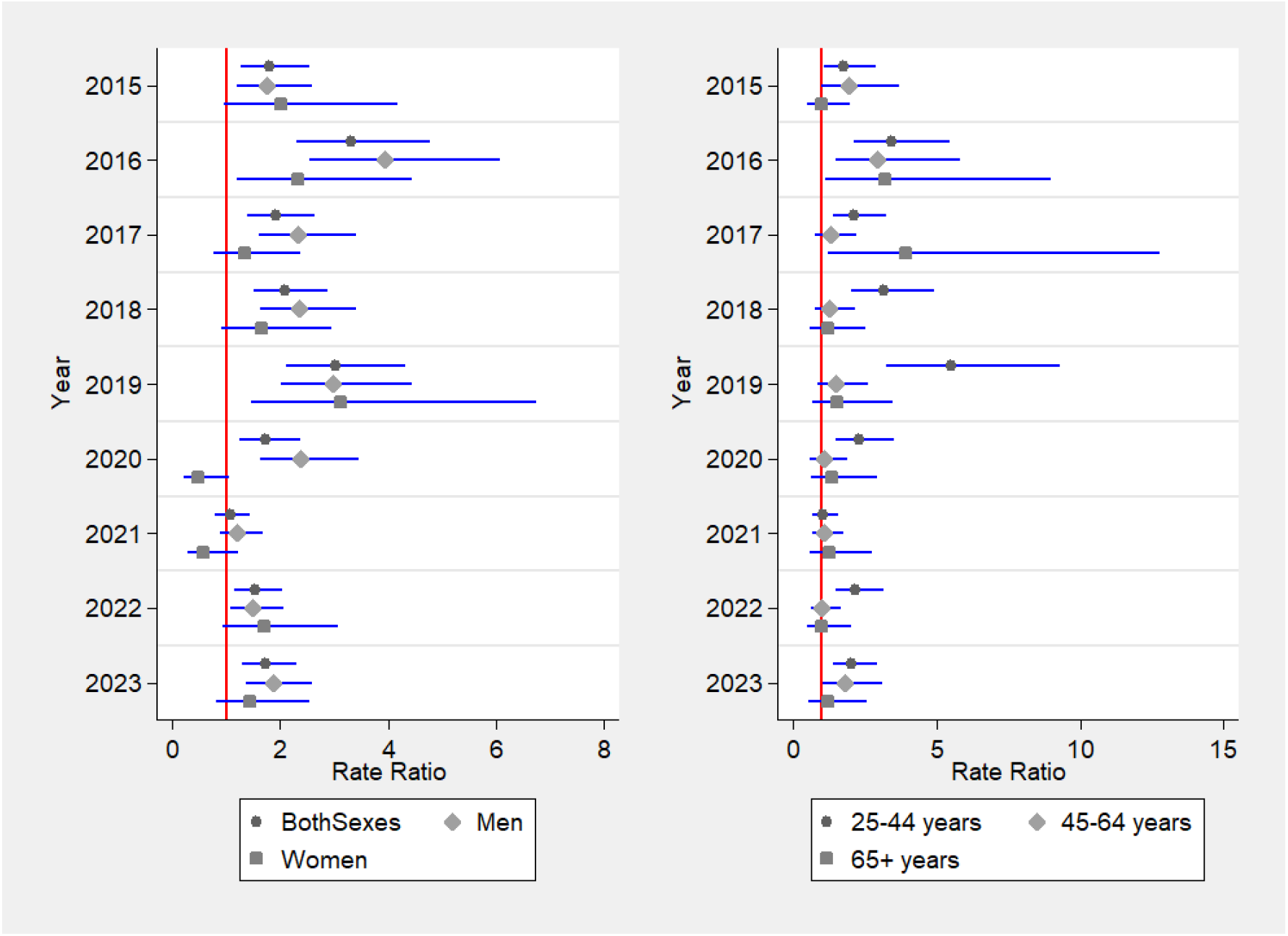
Annual suicide* rate ratios using as standard the population of medium-high educational level. Vertical segments are 95% confidence intervals for the ratio of mortality rates computed with mortality including all explanatory variables and an interaction between year and educational level. * Suicides including deaths of undetermined intent by hanging (ICD-10 code: Y20) and drowning (ICD-10 code: Y21).

**Figure 3A.**
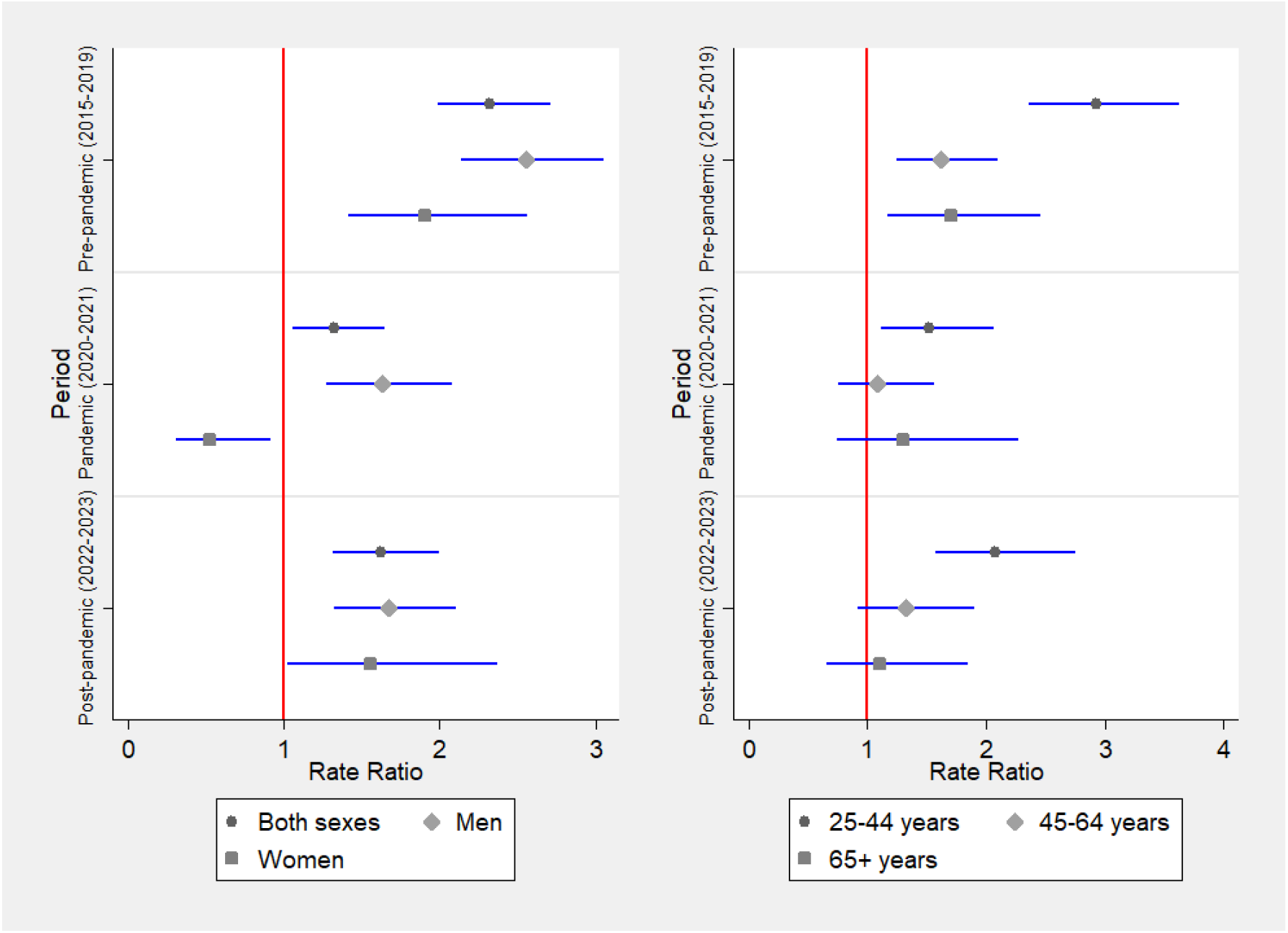
Suicide* rate ratios using as standard the population of medium-high educational level. Vertical segments are 95% confidence intervals for the ratio of mortality rates computed with mortality including all explanatory variables and an interaction between period and educational level. * Suicides including deaths of undetermined intent by hanging (ICD-10 code: Y20) and drowning (ICD-10 code: Y21).

